# Global analysis of daily new COVID-19 cases reveals many static-phase countries including US and UK potentially with unstoppable epidemics

**DOI:** 10.1101/2020.05.08.20095356

**Authors:** Xinmiao Fu

## Abstract

The COVID-19 epidemics are differentially progressing in different countries. Here, comparative analyses of daily new cases reveal that 61 most affected countries can be classified into four types: downward (22), upward (20), static-phase (12) and uncertain ones (7). In particular, the 12 static-phase countries including US and UK are characterized by largely constant numbers of daily new cases in the past over 14 days. Furthermore, these static-phase countries are overall significantly lower in testing density but higher in the level of positive COVID-19 tests than downward countries. These findings suggest that the testing capacity in static-phase countries is lagging behind the spread of the outbreak, i.e., daily new cases (confirmed) are likely less than daily new infections and the remaining undocumented infections are thus still expanding, resulting in unstoppable epidemics. As such, increasing the testing capacity and/or reducing the COVID-19 transmission are urgently needed to stop the severing crisis in static-phase countries.

**Highlights:**

1. 61 most affected countries can be classified into four types: downward, upward, static-phase and uncertain ones.
2. The 12 static-phase countries including US and UK are characterized by largely constant numbers of daily new cases in the past over 14 days.
3. Static-phase countries show lower testing density but higher level of positive COVID-19 tests than downward countries.
4. In static-phase countries, daily new cases (confirmed) are likely less than daily new infections and the remaining undocumented infections are thus still expanding.

## Introduction

The outbreak of 2019 novel coronavirus diseases (COVID-19) has become a pandemic and are devastating more than 150 countries ^1, 2^. As of 23 April 2020, there have been over 2.5 million confirmed COVID-19 cases and 180,000 deaths in the world^1^. While China and South Korea are close to successfully contain the COVID-19 epidemic by mass testing and implementing comprehensive prevention measures ^2^, other countries are in battles of different stages against the crisis. We previously forecasted that Italy and the United States were similar to Hubei, China in the severity of the COVID-epidemic, and the US was even more severe than Italy ^3^. Here we performed analyses on over 60 most affected countries to characterize the progress of the epidemic in each country, which may help to get better preparedness and optimize responses and efforts.

## Methods

### Data source

The data of daily new cases of each country were collected from Worldometer (https://www.worldometers.info/coronavirus/). Levels of daily positive COVID-19 tests in the two most affected US states (New York and New Jersey) were collected from the website of the COVID Tracking Project (https://covidtracking.com/api). The data of daily tests for US, Italy and South Korea were collected from the Website of One World Data (https://ourworldindata.org/grapher/full-list-cumulative-total-tests-per-thousand) and Data for UK from COVID-19 UK UPDATE (https://covid19uk.live/#/).

### Statistical analysis

Statistics were performed using Microcal Origin software with ANOVA algorithm, and significance level was set at a *P* value of 0.05.

## Results

### Identification of static-phase countries with largely constant numbers of daily new cases

We collected the data of daily new cases of the most affected 61 countries (at least with over 1800 total confirmed cases as of 19 April 2020) from the website of Worldometer ^1^ and then compared the trends of daily new cases among them. As summarized in **Table 1**, these 61 countries can be classified into four type, i.e., 22 downward, 20 upward, 12 static-phase and 7 uncertain countries, as described as follows. First, daily new COIVD-19 cases in the 22 downward countries appear to decline in the past over 14 days after passing peaks. These countries include Spain, Italy, Germany and Iran (**Fig. 1A**), China and South Korea that have successfully diminished the domestic spread of the virus (**Fig. S1A**), and many others (**Figs. S1B, S1C** and **S1D**). Iceland, Japan and France somehow show declining trends (**Figs. S1E** and **S1F**) but need more time to be verified. Second, there are 20 upward countries, in which daily new cases are rapidly (**Figs. 1A, S2A** and **S2B**) or slowly (**Figs. S2C**) increasing over time. In particular, Singapore had been believed to contain the epidemic successfully but recently reports drastic increase in new cases (**Fig. 1A**). Third, daily new cases in the 6 uncertain countries vary dramatically such that it is hard to determine the trends of their epidemics (**Figs. S3A** and **S3B**); Turkey seems to stabilize in daily new cases in the past 7 days and needs more time for judgement (**Fig. S3C**).

**Table 1.**
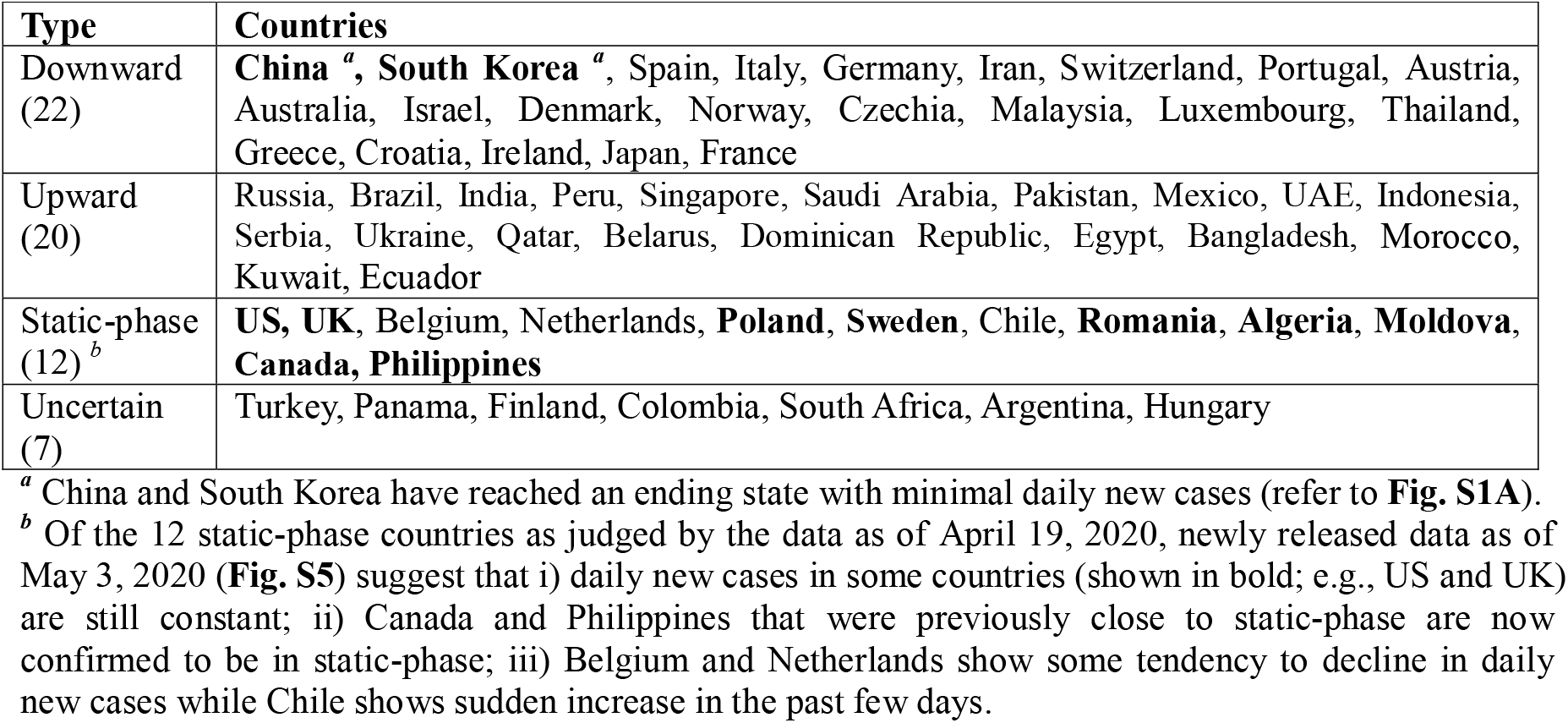
Classification of the 61 most affected countries according to their trends of daily new cases.

| **Type** | **Countries** |
| --- | --- |
| Downward<br>(22) | **China**<sup>a</sup>, **South Korea**<sup>a</sup>, Spain, Italy, Germany, Iran, Switzerland, Portugal, Austria, Australia, Israel, Denmark, Norway, Czechia, Malaysia, Luxembourg, Thailand, Greece, Croatia, Ireland, Japan, France |
| Upward<br>(20) | Russia, Brazil, India, Peru, Singapore, Saudi Arabia, Pakistan, Mexico, UAE, Indonesia, Serbia, Ukraine, Qatar, Belarus, Dominican Republic, Egypt, Bangladesh, Morocco, Kuwait, Ecuador |
| Static-phase<br>(12)<sup>b</sup> | **US, UK**, Belgium, Netherlands, **Poland, Sweden**, Chile, **Romania, Algeria, Moldova, Canada, Philippines** |
| Uncertain<br>(7) | Turkey, Panama, Finland, Colombia, South Africa, Argentina, Hungary |

<sup>a</sup> China and South Korea have reached an ending state with minimal daily new cases (refer to **Fig. S1A**).
<sup>b</sup> Of the 12 static-phase countries as judged by the data as of April 19, 2020, newly released data as of May 3, 2020 (**Fig. S5**) suggest that i) daily new cases in some countries (shown in bold; e.g., US and UK) are still constant; ii) Canada and Philippines that were previously close to static-phase are now confirmed to be in static-phase; iii) Belgium and Netherlands show some tendency to decline in daily new cases while Chile shows sudden increase in the past few days.

**Figure 1.**
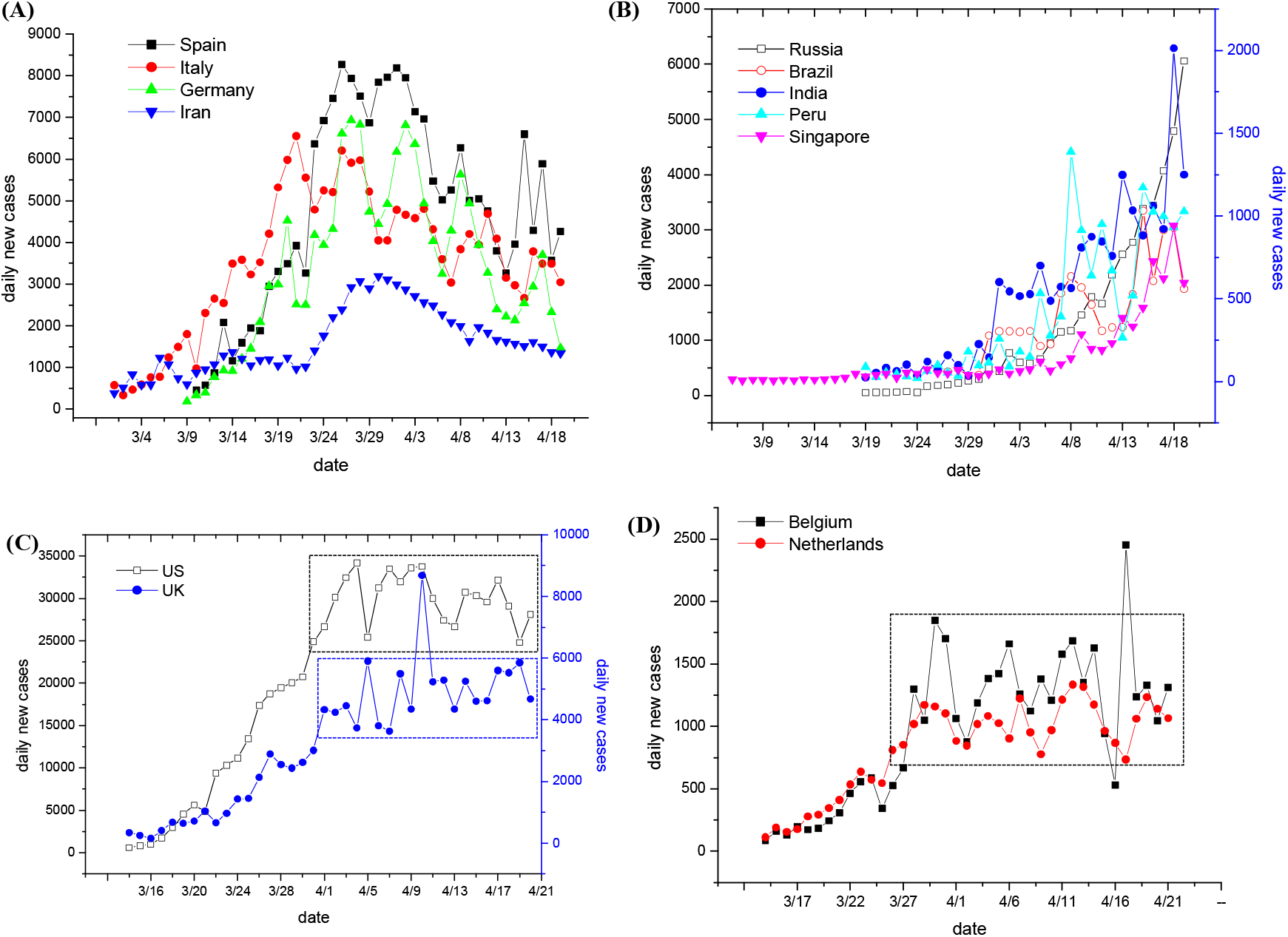
Daily new cases in typical downward, upward and static-phase countries over time. (**A, B, C, D**) Daily new cases in typical downward (panel A), upward (panel B) and static-phase countries (panels C and D). Other countries of each type are shown in Fig. S1, S2 and S4, respectively. Data (as of 19 April, 2020) are collected from Worldometer ^1^. For each country, the starting date was set when the cumulative confirmed cases reached 1% of the current cumulative ones as of 19 April, 2020. Dotted windows in panels C and D show the period (over 14 days) with invariable daily new cases. Results of other countries in these categories are shown in **Figs. S1, S2** and **S4**.

Strikingly, we found that 12 countries including US and UK entered into static states that are characterized by largely constant numbers of daily new cases in the past over 14 days (as indicated by dotted windows in **Figs. 1C, 1D** and **S4A**; for detail, refer to **Table S1**). It has been reported that the incubation period of COVID-19 ranges from 1-14 days with a mean of 5-6 days ^4^, and therefore maintaining invariable daily new cases within a period of over 14 days is unusual. Notably, such static-phase periods for US, UK, Belgium and Netherlands are even more than 20 days (**Figs. 1C** and **1D**; **Table S1**). In addition, Canada and Philippines are close to such static states (**Fig. S4B**), with daily new cases of the former showing slightly increase in the past four days and the latter being varying drastically. During revising and resubmitting the manuscript, we closely monitored the newly released data, which suggest that most of the 12 static-phase countries are still constant in daily new cases as of May 3, 2020 (as shown in bold style, **Table 1**; for detail, refer to **Fig. S5**).

### Static-phase countries show significantly lower testing density but higher level of positive COVID-19 tests than downward countries

To unravel why these static-phase countries show invariably new case, we compared the four types of countries with respect to case density, testing density and the level of positive COVID-19 tests (for detail, refer to **Table S2**). Results indicate that the 12 static-phase countries are overall comparable in the case density with the 22 downward countries (p=0.375; **Fig. 2A**), but they are significantly lower in the testing density (p=0.016; **Fig. 2B**) and higher in the level of positive COVID-19 tests (p=0.028; **Fig. 2C**) than the latter. Consistently, upward countries are significantly lower in case density than downward countries (p=0.003; **Fig. 2A**). These observations raise a possibility that testing capacity may limit the full identification of COVID-19 infected persons in the static-phase countries where relative high levels of infections have been already presented in the community (as indicated by the relatively high level of positive COVID-19 tests; **Fig. 2C**).

**Figure 2.**
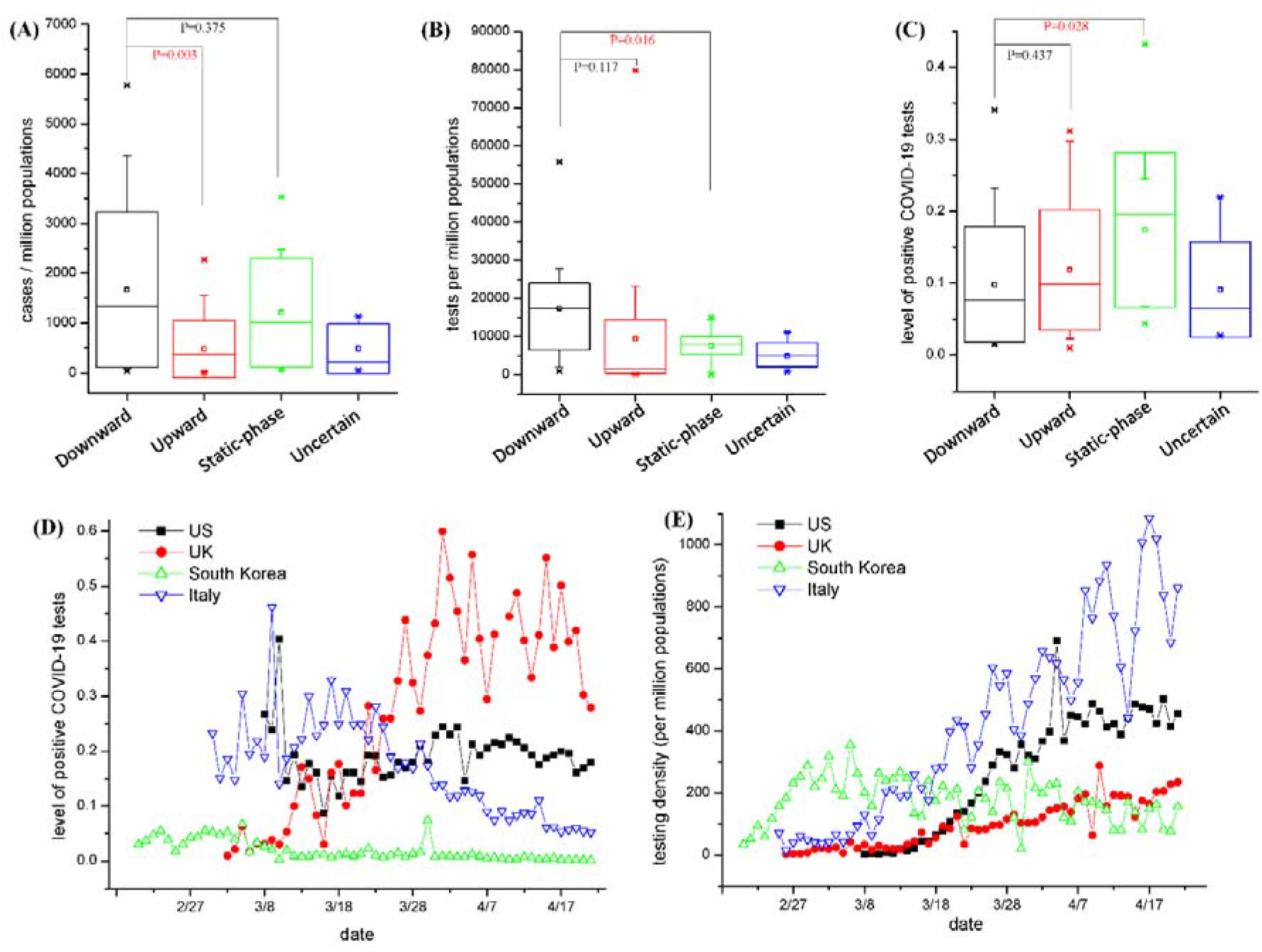
Comparative analyses of COVID-19 tests between downward and static-phase countries. (**A, B, C**) Difference analysis of case density (panel A), testing density (panel B) and level of positive COVID-19 tests (panel C) among the four types of countries. Data of each type of countries were plotted as mean±SD at 95% confidence intervals (in the box), with median being shown as short lines. Statistics were performed using Microcal Origin software with ANOVA algorithm, and significance levels (*P* value) are designated. P values less than 0.05 are colored in red. (**D, E**) Comparisons of time-dependent levels of positive COVID-19 tests (panel A) and testing density (panel B) between typical static-phase countries (US and UK) and downward countries (South Korea and Italy). Data for US, Italy and South Korea were collected from the Website of One World Data (https://ourworldindata.org/grapher/full-list-cumulative-total-tests-per-thousancl) and Data for UK from COVID-19 UK UPDATE (https://covid19uk.live/#/).

### US and UK have lower testing intensity but higher level of positive tests than South Korea and Italy

To further verify the above possibility, we compared the daily testing density between typical static-phase countries (US and UK) and downward countries (South Korea and Italy). Data presented in **Fig. 2D** indicate both US and UK are lower than Italy in the testing density even if the latter has significantly mitigated the outbreaks in the past month (**Fig. 1A**) while the former two are still expecting the appearance of epidemic peaks. Notably, testing density in South Korea has declined since April but is still comparable with that in UK (**Fig. 2D**), although South Korea has approached to the ending stage in the outbreak (**Fig. S1A**). These observations suggest that both US and UK have not yet performed sufficient testing to identify the infected COVID-19 cases in comparison with what Italy and South Korea have done regardless of their epidemic status.

More severely, US and UK have surpassed Italy in the level of positive COVID-19 tests since late March, and in the US the level of positive tests has been close to the ever highest peak in Italy (up to 30%) occurring in mid-March and in UK it is even much higher than the latter (up to 50%; **Fig. 2E**). In particular, the levels of positive tests in New York and New Jersey states, the epicenter of the outbreaks in the US, are even up to 40% and 50%, respectively (**Fig. S6**). These observations implicate that infection levels in the populations of US and UK would be, if not higher, comparable with the highest level of Italy throughout its epidemic. Consistently, the level of positive tests in South Korea is as lower as below 5%, presumably due to the limited spread of the virus in the community and/or mass testing.

## Discussion

In summary, we surprisingly found that among the 61 most affected countries 12 have been in a unique static-phase state with highest throughout the epidemic course but largely invariable daily new cases in the past over 14 days. The infection levels among the populations of these static-phase countries are even higher than the downward countries (**Fig. 2C**), and the absolute testing densities in certain static-phase countries including the US, Belgium, Canada and Netherlands are even higher than some of the downward countries (e.g., Malaysia and Greece) (refer to **Table S2**). It follows that invariable daily new cases in static-phase countries should be conceivably resulted from relative insufficient testing in the context of a large pool of undocumented or unidentified infected cases, which is supported by recent antibody screening in New York ^5^. Along this logic, daily new (confirmed) cases are most likely less than daily new infections in these static-phase countries. As such, the undocumented or unidentified infected cases would increase over time, which further accelerates the spread of the virus and eventually leads to unstoppable epidemics until the virus has spread to the whole populations.

Accordingly, expand of the testing capacity and interruption of transmissions by quarantine measures are critical for containing the COVID-19 outbreaks because the former increases daily new cases and the latter decreases daily new infections. If the current testing density is insufficient regarding the large infection pool among the populations, it is expected that the daily new cases will be proportionally increased if the testing capacity in static-phase countries can be expanded. In this way, more infected persons would be identified and subsequently isolated and quarantined, which would further reduce the spread of the virus among populations and thus decrease total undocumented or unidentified infections. On the other hand, daily new infections are proportional to total active infections and the secondary attack rate that could only be reduced by quarantine measures ^6^. What China ^4, 7^, Italy ^6^ and Germany ^8^ have done for fighting against COVID-19 in these aspects may inform static-phase countries to optimize efforts.

## Data Availability

all data are included in the manuscript

## Acknowledgments

This work is support by the National Natural Science Foundation of China (No. 31972918 and 31770830 to XF). Authors ae grateful to graduate students (Boyan Lv, Zhongyan Li, Zhongyu Chen, Shuang Zhang, Fengqi Sun, Zuqin Zhang, all from Prof. Xinmiao Fu’s research group at Fujian Normal University) for data collection.

